# Exploring the relationship between resting state intra-network connectivity and accelerometer-measured physical activity in pediatric concussion: A cohort study

**DOI:** 10.1101/2021.07.15.21260586

**Authors:** Bhanu Sharma, Joyce Obeid, Carol DeMatteo, Michael D. Noseworthy, Brian W. Timmons

**Affiliations:** Child Health and Exercise Medicine Program, Department of Pediatrics, McMaster University; School of Rehabilitation Science, McMaster University; CanChild Centre for Childhood Disability Research, McMaster University; Imaging Research Centre, St. Joseph’s Healthcare, Hamilton, ON, Canada; Department of Electrical & Computer Engineering, McMaster University; McMaster School of Biomedical Engineering, McMaster University; Department of Radiology, McMaster University

## Abstract

**Objectives:** To explore the association between resting state functional connectivity and accelerometer-measured physical activity in pediatric concussion.

**Methods:** Fourteen children with concussion (aged 14.54 ± 2.39 years, 8 female) were included in this secondary data-analysis. Participants had neuroimaging at 15.3 ± 6.7 days post-injury and subsequently a mean of 11.1 ± 5.0 days of accelerometer data. Intra-network connectivity of the default mode network (DMN), sensorimotor network (SMN), salience network (SN), and fronto-parietal network (FPN) was computed.

**Results:** Per general linear models, only intra-network connectivity of the DMN was associated with habitual physical activity levels. More specifically, increased intra-network connectivity of the DMN was significantly associated with higher levels of subsequent accelerometer-measured light physical activity (F_(2,11)_ = 7.053, p = 0.011, R_a_ ^2^ = 0.562; β = 0.469), moderate physical activity (F_(2,11)_ = 7.053, p = 0.011, R_a_ ^2^ = 0.562; β = 0.725), and vigorous physical activity (F_(2,11)_ = 10.855, p = 0.002, R_a_ ^2^ = 0.664; β = 0.79). Intra-network connectivity of the DMN did not significantly predict sedentary time. Likewise, the SMN, SA, and FPN were not significantly associated with either sedentary time or physical activity.

**Conclusion:** These findings suggest that there is a positive association between the intra-network connectivity of the DMN and device-measured physical activity in children with concussion. Given that DMN impairment can be commonplace following concussion, this may be associated with lower levels of habitual physical activity, which can preclude children from experiencing the symptom-improving benefits of sub-maximal physical activity.

**KEY FINDINGS:** *What are the new findings?:*

- Intra-network connectivity of the default mode network is associated with subsequent accelerometer-measured light, moderate, and vigorous physical activity within the first-month of pediatric concussion
- Similar associations with physical activity are not observed when examining the intra-network connectivity of the sensorimotor network, salience network, or fronto-parietal network
- Improved connectivity within the default mode network may lead to increased participation in light to vigorous physical activity in pediatric concussion

*How might it impact on clinical practice in the future?:*

- Default mode network impairment is commonplace in concussion, and this may limit children from experiencing the symptom-improving benefits of physical activity
- Adjunctive interventions (e.g., mindfulness) that improve the health of the default mode network should be further studied in pediatric concussion

## INTRODUCTION

Concussions are mild traumatic brain injuries that result in functional neurological disturbance in the absence of gross structural damage^1^. While on the mild end of the brain injury spectrum, concussions can nonetheless result in enduring symptoms that negatively impact multiple aspects of daily living^2^. In children, a population in which rates of concussion are increasing rapidly^3-5^, nearly 30% experience symptoms that persist beyond the expected recovery window of four weeks^1,6,7^. These symptoms can lead to quality-of-life deficits that can last for months^8-11^.

A growing body of research has used resting-state functional magnetic resonance imaging (rs-fMR). In this study, rs-fMRI was used to provide insight into areas of the brain that are synchronously active at rest in the absence of any particular cognitive stimuli) to explore the functional neuropathology of concussion. While the brain regions and/or networks studied, time of imaging, and rs-fMRI analysis methods themselves have varied across studies, studies consistently show that there are rs-fMRI disturbances in children with concussion in comparison to their healthy peers^12-23^. Collectively, these studies have shown patterns of both hyper-and hypo-connectivity within the initial weeks of injury^24^, abnormal rs-fMRI activity in children with protracted symptoms^15^, and a persistence of functional impairment in asymptomatic children who have been medically cleared to return-to-sport^21^.

With rs-fMRI disturbances in pediatric concussion now better characterized, studies have aimed at understanding the relationship between functional connectivity and other relevant and widely studied clinical outcomes including symptoms, sleep, cognition, mood^15-17,25^. However, the associations between resting state brain activity and other salient features of pediatric concussion are still understudied. In particular, the impact of rs-fMRI activity on habitual physical activity in pediatric concussion remains unexplored. Within the current landscape of concussion research and clinical management, wherein the importance of exercise has been increasingly recognized and established in recent years^26,27^, this represents a considerable knowledge gap. There is now a shift away from prolonged rest^28,29^, which was the former *status quo* in concussion management^30^.. Conversely, sub-maximal aerobic exercise studies in concussion–and meta-analyses of them–suggest that engaging in physical activity within two-weeks of injury improves symptoms^26,27,31^. Therefore, there is now considerable evidence that exercise, and conversely prolonged rest, have an impact on concussion recovery. What is not known is how the widely established rs-fMRI disturbances observed in pediatric concussion relate to sedentary time or physical activity levels post-injury.

With the recent wave of research on exercise in pediatric concussion, understanding the relationship between functional brain impairment and physical activity has become as germane as understanding the relationship between rs-fMRI and symptomatology or other clinical features. Therefore, in this study, we examined the relationship between intra-network rs-fMRI activity (within four widely researched and validated resting state networks) and accelerometer-measured habitual physical activity and sedentary time up to one-month post-injury in children with concussion. It was hypothesized that reduced intra-network rs-fMRI activity (a measure of functional neuropathology) is associated with increased sedentary time and reduced physical activity levels.

## METHODS

The Hamilton Integrated Research Ethics Board approved this study.

### Design

The data reported here were initially collected as part of the *Back-to-Play* study, a larger cohort study (led by the senior authors) with the goal of informing return-to-activity guidelines for children with concussion. This report is a secondary data analysis of accelerometer and rs-fMRI data collected as part of the larger cohort. The present sample is comprised only of participants from the parent study with both neuroimaging data and subsequently collected accelerometer data (up to 1-month post-injury).

### Participants

Participants in the parent study were recruited at McMaster Children’s Hospital and/or its affiliated rehabilitation and sports medicine clinics. Patients who were diagnosed with concussion by a member of our clinical team were informed about the *Back to Play* study. Those interested in participating were referred to our research team for more information about its objectives, risks, and potential benefits. After this initial discussion, those intent on participating were consented (or assented, along with parental consent, if aged under 16 years) recruited. An intake assessment was then scheduled by the research team as soon after the initial clinical consultation as possible. Exclusion criteria for the larger study included more severe injuries or those requiring more complex care. For the present study, patients were required to have neuroimaging and then subsequently at least 5 days of valid accelerometer wear.

### Data collection procedures

#### Neuroimaging

All neuroimaging data were collected using a 3-Tesla GE Discovery 750 MRI scanner (with a 32-channel phased array receive coil) at the Imaging Research Centre (IRC) at St. Joseph’s Healthcare, Hamilton. A screening questionnaire was performed by the IRC imaging technologist to ensure that the scan could be performed safely, and to inform patients and their families about the MRI procedure. The technologist then positioned the patient in the MRI, immobilizing their head with foam pads to minimize motion and to improve patient comfort.

The neuroimaging battery began with a 3-plane localizer with calibration sequences. Anatomical images were then collected, per a 3D IR-prepped fast SPGR T1-weighted sequence (TR/TE=11/36/4.25ms, flip angle=12°, interpolated 512×512 matrix, 22 cm FOV). Immediately prior to the resting state scan, a fieldmap was acquired (to correct for magnetic field inhomogeneities) using the same geometry as the functional scan (as follows). Resting state functional data were collected using axial 2D acquisition, gradient echo EPI, TR/TE=2000/35ms, flip angle=90°, 64×64 matrix, 300 time points (10min), 22 cm FOV). During the resting state scan, patients were asked to remain awake with their eyes open, and not think of “anything in particular”.

#### Accelerometry

Patients were given a compact and light-weight waist-worn tri-axial accelerometer at their intake assessment to wear until self-reported symptom resolution, at which point the accelerometer was mailed back to the study team and acceleration data were downloaded. The accelerometer used in the *Back-to-Play* study was the ActiGraph GT3x (Pensacola, FL), which has demonstrated high reproducibility in measuring physical activity in acquired brain injury^32^. Per the parent study, movement was recorded continuously at 30-Hz and downloaded into 3-second epochs. Patients were also given a logbook, in which they noted when the device was put on and off in the morning and evening, respectively, as well as any other times the device was removed (when participating in water-based activities, for example). Patients were instructed by the research team at the intake assessment to wear the accelerometer on the right hip during all waking hours, except for water activities, and how to use the logbook.

### Data processing and analyses

#### Neuroimaging

All MRI pre-processing and analyses were performed in CONN 19c^33^, which was run using SPM12 and MATLAB 2020a (Mathworks, Natick, MA). The only exception in the pre-processing pipeline was that functional data were unwarped using the B_0_ maps acquired immediately prior to the rs-fMRI scan outside of CONN 19c using *epiunwarp*^34^, which draws on functionality in FSL^35,36^. The B_0_ -corrected maps were then uploaded into CONN 19c along with respective anatomical data for pre-processing.

The following steps were involved with pre-processing, and were guided by recommendations within CONN 19c and associated publications^37,38^. First, functional data were realigned and co-registered to a reference image (and adjusted to the movement-defined deformation field associated with the reference image). Second, slice-timing correction was performed to time-shift and resample the rs-fMRI data to coincide with the mid-point of each TR. Third, using SPM’s Artifact Detection Tool (ART)^39^, outlier scans in the functional data were flagged based on subject-motion exceeding a 0.9mm framewise displacement or BOLD signal fluctuations > 5 standard deviations from the mean signal. Fourth, functional data are normalized/registered to MNI space, based on posterior tissue probability maps. Fifth, spatial smoothing was performed, with a Gaussian kernel of full-width at half-maximum of 6mm. Subsequently, de-noising was performed in CONN 19c which involved two steps: 1) an anatomical component-based noise correction procedure (`aCompCorr`) to “regress out” noise components associated with cerebral white matter/cerebrospinal regions, outlier scans, subject motion (based on a 12-parameter affine transformation of the anatomical to functional data)^40-42^, and 2) temporal filtering, using a filter ranging from 0.008Hz and above 0.01Hz. After both pre-processing and de-noising, data were inspected visually and per the quality assurance metrics in CONN 19c.

Once data were ready for analysis, intra-network connectivity of four widely studied and validated brain networks was computed^43,44^, namely the default mode network (DMN), sensorimotor network (SMN), salience network (SN) and fronto-parietal network (FPN) and habitual physical activity in children with concussion. Intra-network connectivity was selected as the measure of choice for this preliminary investigation as it is representative of a resting state network holistically, as has been shown in publications related to other neurological populations^45-48^.

Average within-network connectivity were computed using the Matlab command line script `conn_withinbetweenROItest`, which extracted and exported single-subject intra-network correlation values which were Fisher Z-transformed to improve normality. Seed regions are listed in **Table 1**.

**Table 1:** Seed regions used to calculate the intra-network connectivity of the four resting state networks of interest.

| Network | Seeds |
| --- | --- |
|  | MNI co-ordinates (x, y, z) |
| DMN | Medial pre-frontal cortex<br>(1, 55, -3) |
|  | Lateral parietal, left<br>(-39, -77, 33) |
|  | Lateral parietal, right<br>(47, -67, 29) |
|  | Posterior cingulate cortex<br>(1, -61, 38) |
| SMN | Lateral seed, left<br>(-55, -12, 29) |
|  | Lateral seed, right<br>(56, -10, 29) |
|  | Superior seed<br>(0, -31, 67) |
| SN | Anterior cingulate cortex<br>(0, 22, 35) |
|  | Anterior insula, left<br>(-44, 13, 1) |
|  | Anterior insula, right<br>(47, 14, 0) |
|  | Rostral pre-frontal cortex, left<br>(-32, 45, 27) |
|  | Rostral pre-frontal cortex, right<br>(32, 46, 27) |
|  | Supramarginal gyrus, left<br>(-60, -39, 31) |
|  | Supramarginal gyrus, right<br>(62, -35, 32) |
| FPN | Lateral pre-frontal cortex, left<br>(-43, 33, 28) |
|  | Lateral pre-frontal cortex, right |
|  | (41, 38, 30) |
|  | Posterior cingulate cortex, left |
|  | (-46, -58, 49) |
|  | Posterior cingulate cortex, right |
|  | (52, -52, 45) |

#### Accelerometry

Data were downloaded from the ActiGraph GT3x devices using the accompanying software package, ActiLife Version 6 (Pensicola, FL). Triaxial accelerations collected at 30-Hz were downloaded into 3-sec sampling intervals or epochs, which were selected to reflect the median bout duration for high-intensity activities in children^49^. Once downloaded and converted into a 3-second epoch, wear-time validation was performed in ActiLife. First, a semi-automated procedure was performed to define a non-wear period of minimum length of 5-minutes. These periods were then inspected against the on-and off-times recorded in the patient logbooks, with non-wear periods excluded if they matched records from the patient log books. Days of accelerometer data without an associated log book entry were excluded. Data were then scored to determine activity intensity according to widely used Evenson cut-points^50^ which have the following activity count ranges: sedentary time (0-25 counts/15s), light physical activity (LPA; 26-573 counts/15s), moderate physical activity (MPA; 574-1002 counts/15s), and vigorous physical activity (VPA; 1003+ counts/15s).

Cleaned and scored data were then exported to SPSS Version 27 (IBM Corp. Released 2020. IBM SPSS Statistics for Windows, Version 27.0. Armonk, NY). Only children with more than 4-wear days of data, with each day comprised of more than 600 minutes of wear time, were included in the analysis^51^. Any accelerometer data beyond 30-days post-injury were excluded from the current analysis, given that the scope of the study.

### Statistical analyses

Average daily sedentary time, as well as average daily LPA, MPA, and VPA were calculated for cleaned and validated accelerometer cases in SPSS. These summary data were then imported into CONN 19c, wherein the *Calculator* tool was used to build a general linear model (GLM) with intra-network connectivity and age as predictors of average daily activity (of the various intensities) or sedentary time. Therefore, for each network of interest, four models were built to examine the association between intra002Dnetwork rs-fMRI connectivity and average time per day in: LPA, MPA, VPA, and sedentary. Given the sample size of our study, additional variables were not included in the model; age was selectively chosen given that connectivity of resting state networks has been shown to vary with age^52^.

## RESULTS

### Overview

Fourteen children with concussion (aged 14.54 ± 2.39 years, 8 female) were imaged at 15.3 ± 6.7 days post-injury and had an average of 11.1 ± 5.0 days of accelerometer data (with ≥ 600 minutes of wear-time per day) thereafter. Average daily monitoring time, sedentary time, and time spent in activity is summarized in **Table 2**.

**Table 2:** Average daily (in minutes/day) monitoring time and activity by intensity in our cohort. (LPA = light physical activity, MPA = moderate physical activity, VPA = vigorous physical activity).

|  | **Mean** | **SD** | **Range** |
| --- | --- | --- | --- |
| **Monitoring time** | 798.3 | 68.0 | 686.8-914.5 |
| **Sedentary** | 631.0 | 75.9 | 504.2-733.2 |
| **LPA** | 116.1 | 33.9 | 68.9-172.2 |
| **MPA** | 28.6 | 11.3 | 13.4-48.6 |
| **VPA** | 22.7 | 16.9 | 3.3-57.8 |

The groupwise average intra-network connectivity values for each of the networks studied are presented in **Table 3** with associated standard deviations and ranges.

**Table 3:** Average intra-network connectivity across the four networks of study (as Fisher Z-transformed correlation coefficients). (DMN = default mode network, SMN = sensorimotor network, SN = salience network, and FPN = fronto-parietal network).

|  | **Mean** | **SD** | **Range** |
| --- | --- | --- | --- |
| **DMN** | 0.50 | 0.15 | 0.21-0.89 |
| **SMN** | 0.60 | 0.21 | 0.18-1.06 |
| **SA** | 0.48 | 0.11 | 0.23-0.70 |
| **FPC** | 0.58 | 0.16 | 0.34-0.81 |

### Relating brain activity and physical activity

From the series of GLMs, the only significant models (and the only network-related beta-coefficients that were significant) pertained to models including the DMN. The intra-network connectivity of the SMN, SA, and FPN did not predict accelerometer-measured sedentary time, LPA, MPA, or VPA. **Figure 1** shows the model-predicted activity levels (along with 95% confidence intervals) by activity type against intra-network connectivity. The figures show that with respect to the DMN, increased intra-network connectivity is closely associated with increased levels of activity, in particular MPA and VPA. Intra-network connectivity of the DMN was not, however, associated with average daily sedentary time.

**Figure 1:**
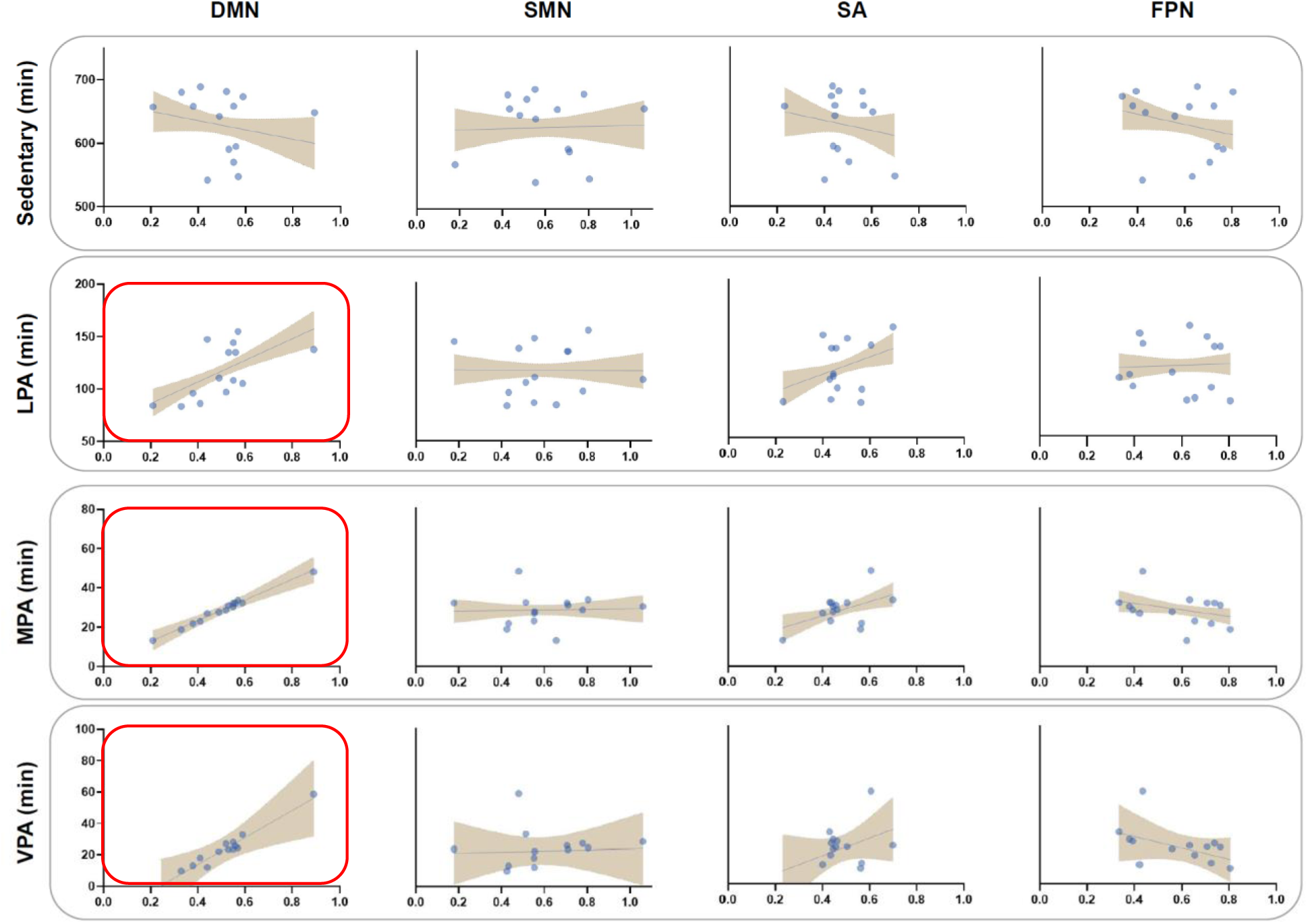
Model-predicted activity levels (in minutes/day) plotted against intra-network connectivity (Fisher Z-transformed correlation coefficients) and a linear fitted line. Only the models highlighted with a red box were significant. (LPA = light physical activity, MPA = moderate physical activity, VPA = vigorous physical activity).

The DMN-specific models and their relevant parameters for all intensities of activity are summarized in **Table 4**. Both unstandardized beta coefficients (B) and standardized beta coefficients (β) are provided for interpretation. Overall, there was a trend for decreased DMN intra-network connectivity predicting more sedentary time, and conversely, increased intra-network connectivity of the DMN was significantly associated with increased LPA, MPA, and VPA. For significant models, the residuals were normally distributed.

**Table 4:** General linear model parameters for DMN-specific analyses. B=unstandardized beta coefficient; β=standardized beta coefficient. (LPA = light physical activity, MPA = moderate physical activity, VPA = vigorous physical activity).

| Sedentary time |  |  |  |  |  |
| --- | --- | --- | --- | --- | --- |
| Predictor | B | 95% CI for B | $\beta$ | t | p |
| DMN | -71.633 | -311.907, 168.640 | -0.146 | -0.656 | 0.525 |
| Age | 20.998 | 5.601, 36.395 | 0.667 | 3.002 | 0.012 |
| Model | $F_{(2, 11)} = 3.653, p = 0.061, R_a^2 = 0.458$ | | | | |
| LPA |  |  |  |  |  |
| Predictor | B | 95% CI for B | $\beta$ | t | p |
| DMN | 102.900 | 86.319, 289.261 | 0.469 | 2.346 | 0.039 |
| Age | -8.486 | -14.671, -2.301 | -0.603 | -3.020 | 0.012 |
| Model | $F_{(2, 11)} = 7.053, p = 0.011, R_a^2 = 0.562$ | | | | |
| MPA |  |  |  |  |  |
| Predictor | B | 95% CI for B | $\beta$ | t | p |
| DMN | 52.908 | 19.627, 86.189 | 0.725 | 3.499 | 0.005 |
| Age | -0.394 | -2.527, 1.739 | -0.084 | -0.407 | 0.692 |
| Model | $F_{(2, 11)} = 6.159, p = 0.016, R_a^2 = 0.528$ | | | | |
| VPA |  |  |  |  |  |
| Predictor | B | 95% CI for B | $\beta$ | t | p |
| DMN | 88.755 | 44.565, 128.945 | 0.792 | 4.526 | 0.001 |
| Age | 1.148 | -1.555, 3.852 | 0.164 | 0.935 | 0.370 |
| Model | $F_{(2, 11)} = 10.855, p = 0.002, R_a^2 = 0.664$ | | | | |

## DISCUSSION

This is the first study to examine the relationship between rs-fMRI impairment and subsequent activity levels in pediatric–or adult–concussion. We demonstrate that intra-network connectivity within the DMN–but not the SMN, SA, or FPN–is associated with accelerometer-measured LPA, MPA, and VPA within the first month of injury in pediatric concussion. Adding to the literature demonstrating associations between DMN impairment and concussion symptoms, depression, anxiety, and sleep-impairment^15,16^, we show that intra-network connectivity within DMN may be implicated with habitual physical activity within the first month of injury in children.

### The DMN and physical activity

Originally, the DMN was considered to be a “day dreaming network”, active in the absence of external stimuli as a type of neural baseline^53^. However, more recent research on the DMN has shown that it is also associated with higher-order cognitive processes and can be active during goal-oriented thoughts and tasks. More specifically, studies now show that the DMN can be engaged when individuals imagine or plan for the future^54^. Other studies suggest that the DMN is important in cognitive processes that result in immediate action or interfere with present behavioural goals^55^. In adults, the DMN has also been linked to “internal mentation”, or introspective thoughts about constructs such as the future or personal intentions^56,57^. Together, these findings would suggest that DMN impairments may have the capacity to influence planned behaviours, such as participation in physical activity, and that DMN impairments can alter how information about physical activity behaviours are perceived, processed, and planned. This would help explain our findings, wherein only the DMN was found to be associated with subsequent levels of physical activity. Further research is needed to build on this possibility. Prior research shows that in young adults with concussion, in comparison to healthy controls, a sub-maximal aerobic fitness test resulted in functional disturbance of the DMN^58,59^. More specifically, said test reduced inter-network connectivity between seed regions of the DMN, namely the PCC and the lateral parietal ROI, PCC and the right lateral parietal ROI, and the PCC and MPFC^59^. Adding to this, our study suggests a bi-directional relationship, wherein intra-network impairment of the DMN may have an impact subsequent physical activity in children with concussion. Given that many studies have demonstrated DMN impairment in pediatric concussion^14-19,21,22^, and our recent work demonstrates that children with concussion are less active than their healthy peers, the current findings suggest that widespread DMN impairment is associated with reduced levels of activity in children with concussion. Larger pediatric concussion cohorts are required to more definitively characterize the relationship between resting state brain activity and physical activity.

### Exercise and concussion management

The status quo in concussion management is changing. The traditional “rest-is-best” approach is being supplanted by an “exercise is medicine” mindset^30^. However, the present study suggests that in pediatric concussion, DMN impairment (which is a common in concussion) has a moderate association with physical activity. This underscores the need for physicians to actively advise pediatric concussion patients to engage in safe, sub-maximal aerobic exercise after a short (24-48 hour) period of rest, as suggested by current guidelines^1^. Otherwise, patients with DMN impairment may not be physically active, and this precludes them from experiencing the benefits of light physical activity on concussion symptoms. Further, in the ultimate interest of promoting physical activity, interventions such mindfulness can be prescribed acutely given their positive impact on the functional connectivity of the DMN^60-64^. Mindfulness is not a contraindication to any other medications that may be prescribed after concussion, and it has other established benefits in brain injury, including improved self-efficacy^65,66^. Future research is needed in this area.

### Generalizability

Our findings need to be studied in adults to understand if they generalize outside of a pediatric context, as several factors may make our findings pediatric-specific. For example, DMN functional connectivity is age-dependent, peaking in adulthood while being less coherent during childhood and senescence^52^; physical activity also decreases with age^67^ This would suggest that the association between intra-network functional connectivity of the DMN and physical activity may be variable in children when compared to adults.. Further, cardiorespiratory fitness is also related to the functional integrity of multiple brain networks, including the DMN^68^. While the present study did not control for cardiorespiratory fitness (though all participants had sport-related injuries and were physically active prior to injury), similar studies in adults should control for this metric given that it declines non-linearly and in an age-dependent rate in adults^69^, which may be confounding in adult samples with large age variance.

## LIMITATIONS

This secondary data analysis is limited by the sample size. Larger studies examining the relationship between the DMN and post-concussion physical activity are warranted. We also cannot rule out the possibility that greater DMN impairment is associated with more severe symptoms that may, in turn, reduce participation in physical activity. This study design does not permit inference on causality. Further, this study did not exhaustively study all resting state networks and/or anatomical regions; future research should expand on the regions of interest. There are also other rs-fMRI and accelerometer analysis methods that can be used to characterize the relationship between brain activity and physical activity. The impact of physical activity on resting state networks in pediatric concussion also warrants study.

## CONCLUSIONS

This exploratory study is the first to examine the association between intra-network connectivity and accelerometer-measured physical activity in pediatric concussion. We found that intra-network connectivity of the DMN–but not the SMN, SA, or FPN–was significantly associated with levels of LPA, MPA, and VPA performed within the first month after pediatric concussion. Given the increasing role of aerobic exercise in concussion management, and that the DMN is commonly perturbed following injury, children with concussion may be less likely to be physically active. Clinical management should continue to encourage participation in sub-maximal aerobic activity in the acute stages of injury.

## Data Availability

Data are available upon reasonable request.

## STATEMENTS

### Competing interests

The authors have no competing interests to declare.

### Contributorship

BW Timmons, MD Noseworthy, and C DeMatteo conceptualized and designed the parent study. C DeMatteo had general oversight over the parent study, with J Obeid and MD Noseworthy maintaining oversight over accelerometer and neuroimaging data collection, respectively. B Sharma created the research question, cleaned and analyzed all accelerometer and neuroimaging data, and prepared the manuscript draft and subsequent revisions.

## Acknowledgements

The study team would like to thank all research participants for their time and interest in our research.

## Funding, grant, and award info

This research was supported by the Canadian Institutes of Health Research (#31257), as well as Doctoral support to B Sharma from the Canadian Institutes of Health Research (CIHR-CGS-D, #157864). BW Timmons is the Canada Research Chair in Child Health & Exercise Medicine.

## Ethical approval information

The study was approved by the Hamilton Integrated Research Ethics Board (www.hireb.ca) # 14-376.

## Data sharing agreement

Data are available upon request.

## Patient involvement

Given that this study is a secondary data analysis, patient and public involvement was not part of this analysis.

## Notes

### Competing Interest Statement

The authors have declared no competing interest.

### Author Declarations

The study was approved by the Hamilton Integrated Research Ethics Board (www.hireb.ca) # 14-376.

